# Computed Tomography Features of COVID-19 in Children: A Systematic Review and Meta-analysis

**DOI:** 10.1101/2020.09.02.20187187

**Authors:** Ji-gan Wang, Yu-fang Mo, Yu-heng Su, Li-chuang Wang, Guang-bing Liu, Meng-Li, Qian-qiu Qin

## Abstract

**Objectives:** To systematically analyze the chest CT imaging features of children with COVID-19 and provide references for clinical practice.

**Methods:** We searched PubMed, Web of Science, and Embase; data published by Johns Hopkins University; and Chinese databases CNKI, Wanfang, and Chongqing Weipu. Reports on chest CT imaging features of children with COVID-19 from January 1, 2020, to August 10, 2020, were analyzed retrospectively and a meta-analysis carried out using Stata12.0 software.

**Results:** Thirty-seven articles (1747 children) were included in this study. The overall rate of abnormal lung CT findings was 63.2% (95% confidence interval [CI]: 55.8–70.6%), with a rate of 61.0% (95% CI: 50.8–71.2%) in China and 67.8% (95% CI: 57.1–78.4%) in the rest of the world in the subgroup analysis. The incidence of ground-glass opacities was 39.5% (95% CI: 30.7–48.3%), multiple lung lobe lesions 65.1% (95% CI: 55.1–67.9%), and bilateral lung lesions 61.5% (95% CI: 58.8–72.2%). Other imaging features included nodules (25.7%), patchy shadows (36.8%), halo sign(24.8%), consolidation (24.1%), air bronchogram signs (11.2%), cord-like shadows (9.7%), crazy-paving pattern (6.1%), and pleural effusion (9.1%). Two articles reported three cases of white lung, another reported two cases of pneumothorax, and another one case of bullae.

**CONCLUSION:** The lung CT results of children with COVID-19 are usually normal or slightly atypica, with a low sensitivity and specificity compared with that in adults. The lung lesions of COVID-19 pediatric patients mostly involve both lungs or multiple lobes, and the common manifestations are patchy shadows, ground-glass opacities, consolidation, partial air bronchogram signs, nodules, and halo signs; white lung, pleural effusion, and paving stone signs are rare.

**CLINICAL IMPACT:** Therefore, chest CT has limited value as a screening tool for children with COVID-19 and can only be used as an auxiliary assessment tool.

*Registration:* This systematic review and meta-analysis was registered in the Prospero International Prospective Register of Systemic Reviews (CRD42020196602).

*Strengths and limitations of this study:* The lung CT findings of children with COVID-19 are usually normal or slightly atypical, with a low sensitivity and specificity compared with that in adults. From a systematic review of current literature, the overall rate of abnormal lung CT findings in children was revealed to be 63.2%. Chest CT has limited value as a screening tool for children with COVID-19 and can only be used as an auxiliary assessment tool. The sample size of some included studies is small, which may affect the results.

## 1. Introduction

In January 2020, severe acute respiratory syndrome-coronavirus-2 (SARS-CoV-2) was identified as the cause of a series of pneumonia cases first diagnosed in Wuhan, Hubei Province, China[1]. Soon, SARS-CoV-2 spread all over the world[2]. As of March, the spread of coronavirus disease 2019 (COVID-19) was recognized as a pandemic by the World Health Organization[3]. In the early stages of the pandemic, it was thought that children were not easily infected [4], but as the pandemic progressed, the number of pediatric cases has gradually increased. Many infected children are asymptomatic, but some patients have fever, dry cough, and fatigue, while others have gastrointestinal symptoms, including abdominal discomfort, nausea, vomiting, abdominal pain, and diarrhea [5]. Computed tomography (CT) is a sensitive tool for diagnosing symptomatic COVID-19 patients. In adult patients, the most common CT manifestation is ground-glass opacities. Other CT manifestations, such as air bronchography, lymph node enlargement, and effusion, are less common [6]. Among the 1014 hospitalized patients with obvious symptoms from Wuhan, China, the CT scans of most patients were abnormal; however, with a sensitivity of 97% and specificity of 25%, the false positive rate was very high [7]. Compared with adults, children have relatively mild symptoms, so CT is not very typical [8]. However, there are few reports on pediatric CT features, and most reports involve small sample sizes. Larger pediatric cohort studies have not been comprehensive enough [9]. Therefore, the lung CT features of children with COVID-19 has been reviewed systematically in this study.

## 1. Evidence Acquisition

### 2.1 Literature Search Strategy

A literature search was conducted through PubMed, Web of Science, Embase, Johns Hopkins University published data, as well as the Chinese databases CNKI, Wanfang, and Chongqing Weipu between January 1, 2020 and August 10, 2020 to collect reports on the characteristics of chest CT of children with COVID-19. Concurrently, online database and manual retrieval were used, and the references included in the literature were traced. Subject-specific and free words were used in the retrieval, and adjustments were made according to the characteristics of the different databases without limitations to language, race, or region. The following search terms were used: “children,” “child,” “kid,” “pediatric”in association with “clinical feature” OR “epidemiology” OR “Imaging” OR “CT” and “2019-nCoV” OR “COVID-19” OR “SARS-CoV-2” OR “Corona Virus Disease 2019.”

### 2.2 Literature Screening and Data Extraction

Two researchers independently searched and screened the articles and collected and cross-checked the data. If there was any dispute, it was resolved by a third researcher.

The inclusion criteria consisted of the following: ① research types: cohort study, case-control study, and case analysis; ② subjects: children with COVID-19; and ③ observation index: imaging features of lung CT or HRCT(High Resolution CT), including lesion distribution, shape, and density change; and accompanying signs.

The exclusion criteria consisted of the following: ① repeated publications of the same research; ② short case reports; and ③ incomplete or missing data analysis, without free or easy access to the data.

### 2.3 Quality Evaluation of the Included Studies

This was a case series study that adhered to the National institute for Clinical excellence (NICE) guidelines for quality evaluation [9]. The evaluation items were as follows: ① cases in the case series came from medical institutions at different levels and from various difference research centers; (2) the research hypothesis or purpose was clearly described; ③ clear reports were included in the exclusion criteria; ④ measurement results were clearly defined; ⑤ the collected data achieved the expected purpose; ⑥ the patient recruitment period was clearly defined; ⑦ the main findings were clearly described; and ⑧ results were analyzed and reported in layers. One point was awarded for each item (maximum 8 points), with a total score ≥4 indicating high-quality research [10]. Two researchers independently evaluated the quality and cross-checked the results.

### 2.4 Statistical Analysis

Meta-analysis was performed using Stata12.0 software (STATA Corp., College Station, TX, USA). First, the original ratio (R) was transformed by double arcsine to conform to a normal distribution, and then the transformed ratio (TR) was analyzed by meta-analysis. The final rate (r) and its 95% confidence interval (CI) were finally obtained by converting the results using the formula: R = (sin(tr/2))^2^. Meta-analysis was carried out using a random-effect model for all studies. The existence of publication bias was judged using the funnel chart, and the significance level was set as α = 0.05.

### 2.5 Patient and public involvement

Patients and public were not involved in this study.

## Results

### 3.1 Literature Screening Process and Results

A total of 2042 related articles were obtained, which were screened layer by layer. Ultimately, 37 studies were included in this report [11–47], including 1747 children with COVID-19. Fig. 1 shows the process and results of the literature screening.

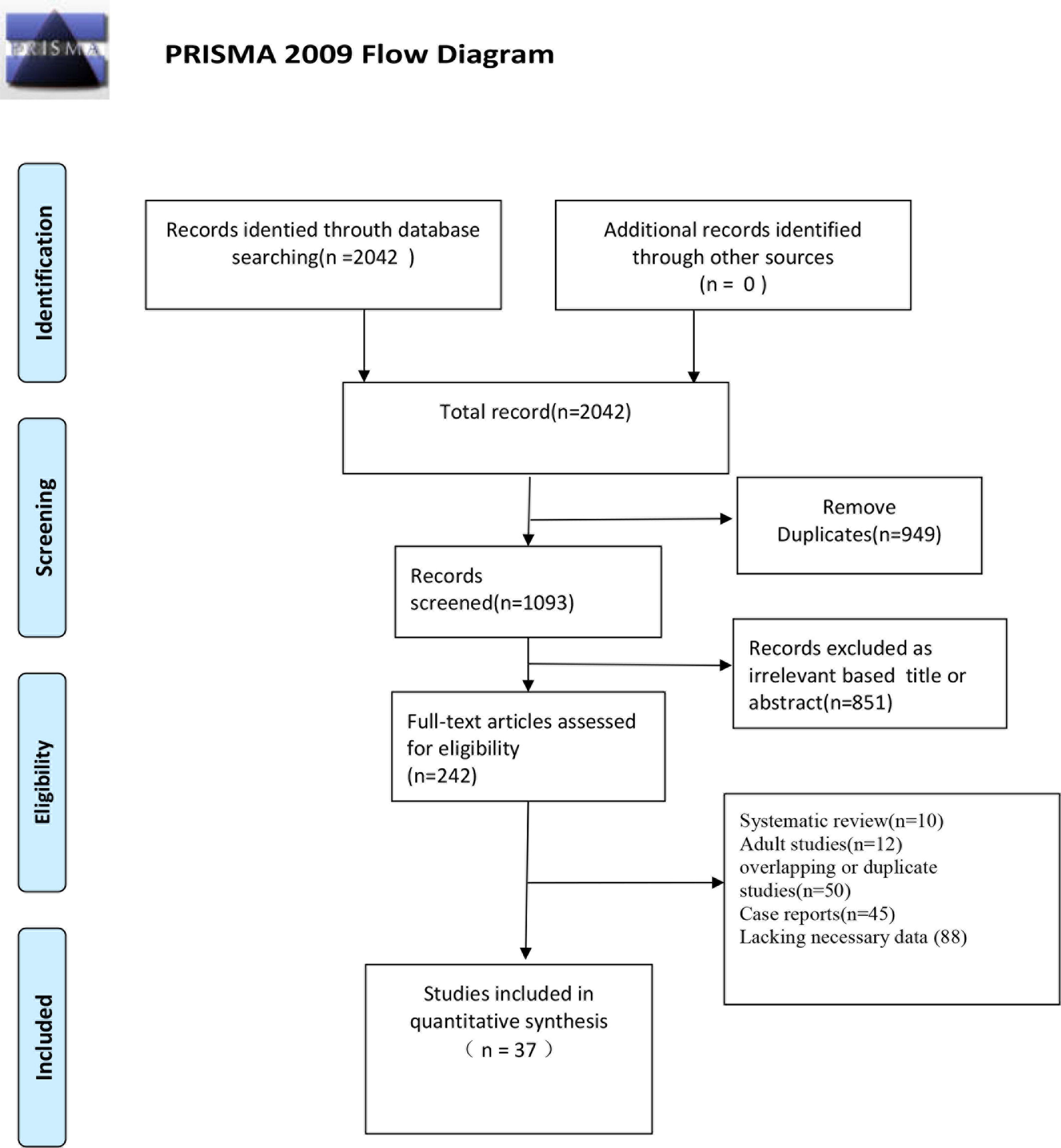

### 3.2 Basic Characteristics and Quality Evaluation Results of the Included Studies

A total of 37 studies [11–47] were included (11 studies from outside China and 26 studies from China). The included studies were published from January 1, 2020, to August 10, 2020. The quality scores of the included studies ranged from 4–8 points, indicating that all were high-quality studies (≥4 points). Table 1 summarizes the basic characteristics of the included studies.

**Table 1:**
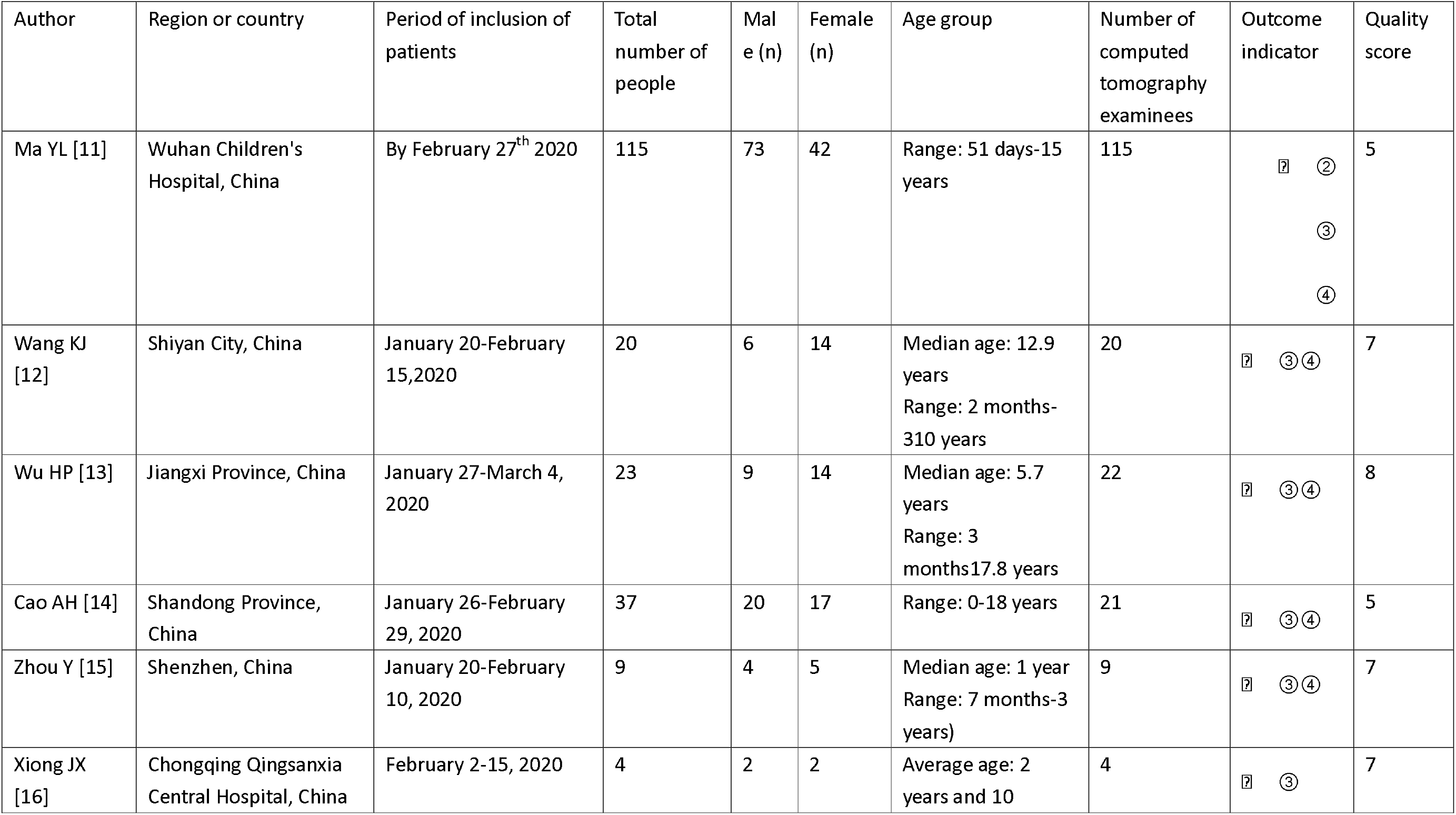
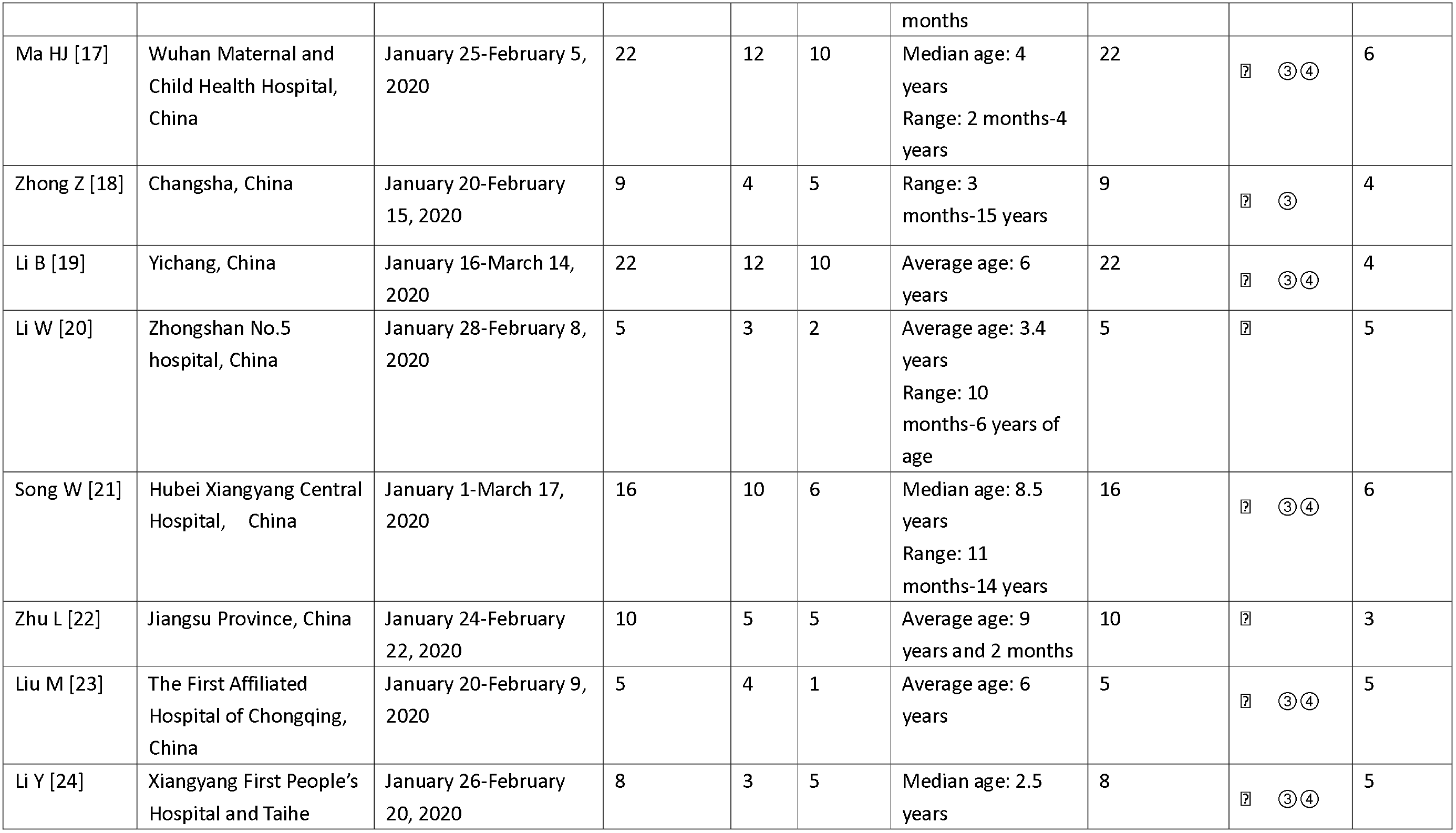
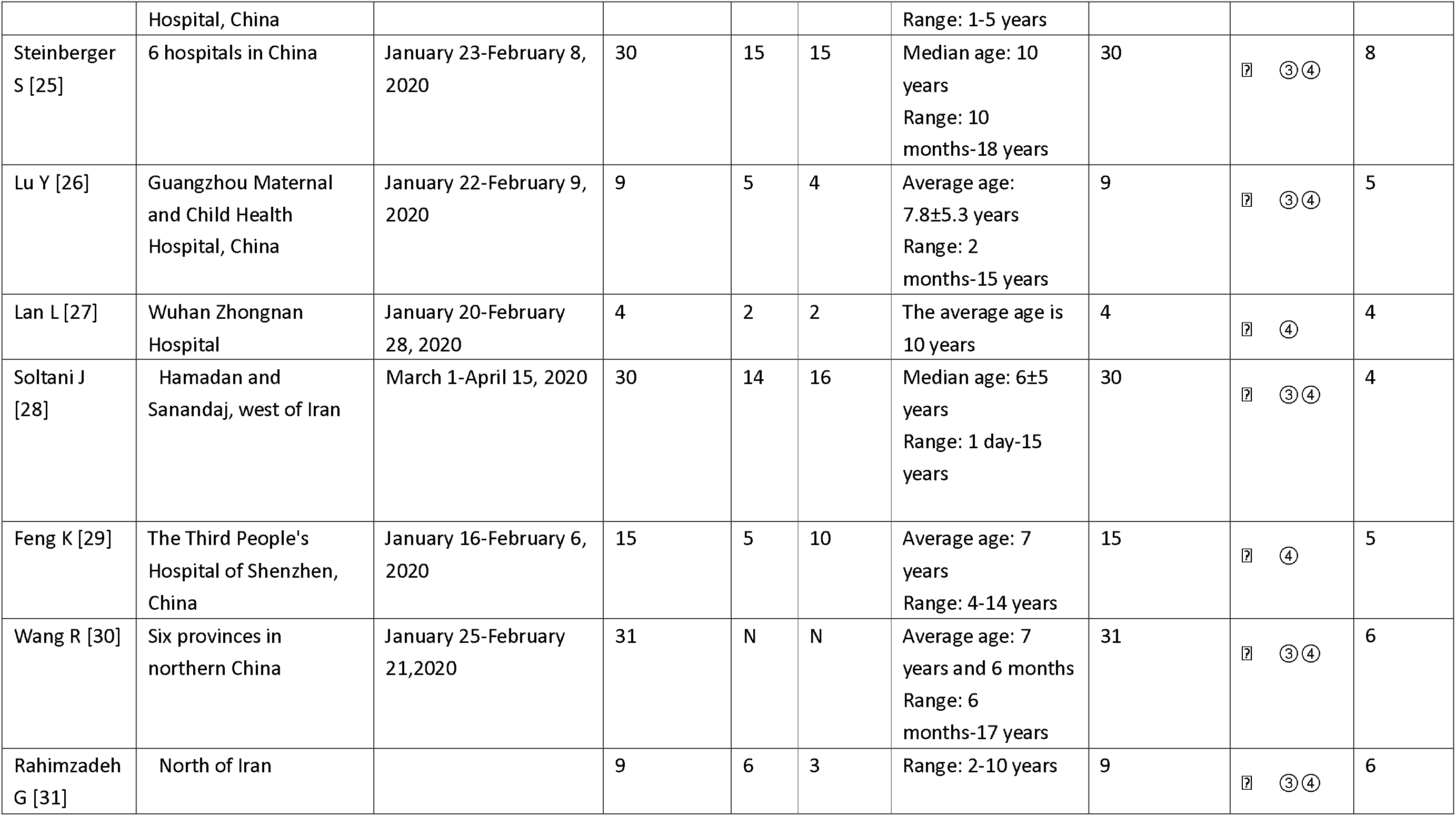
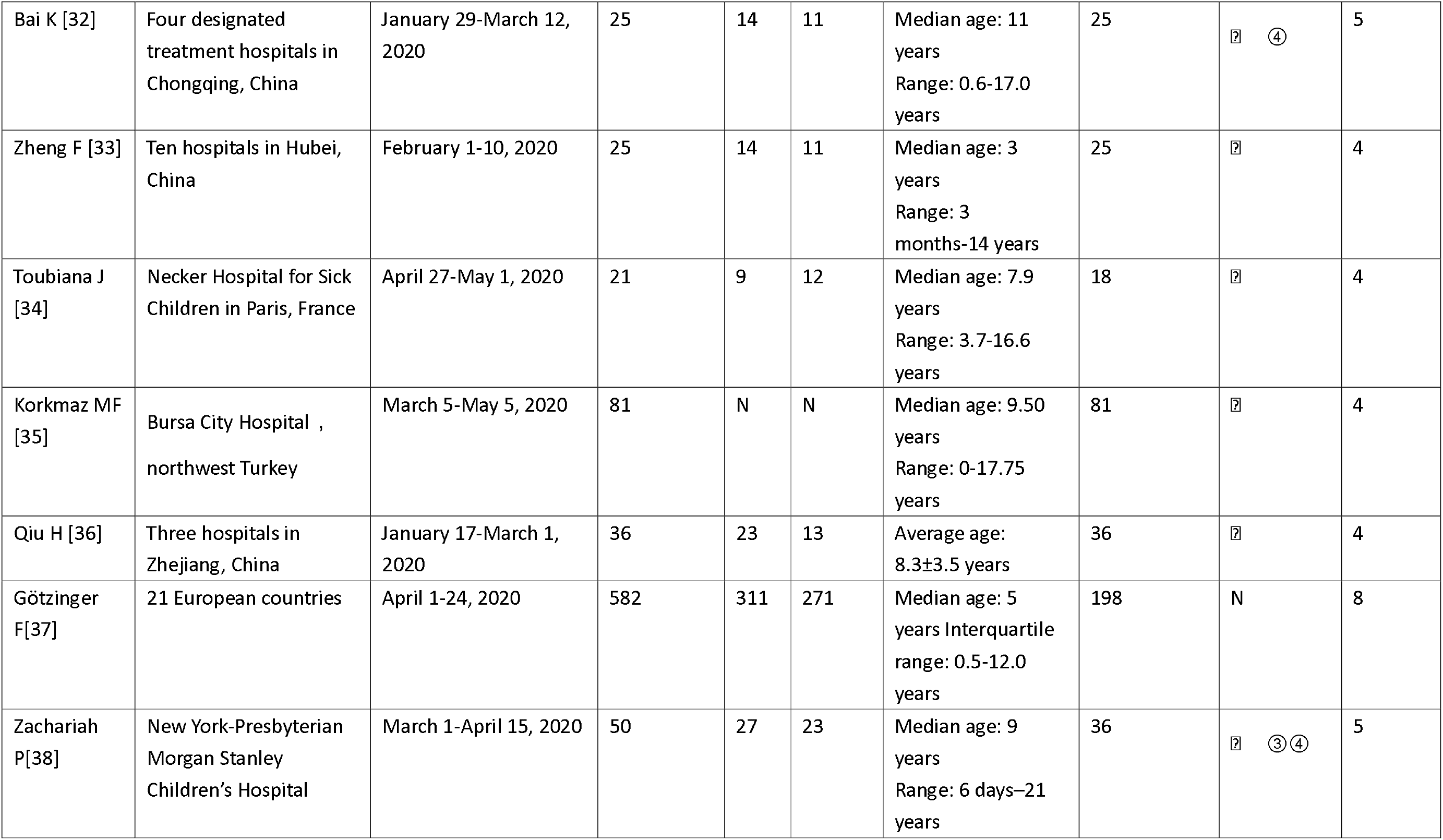
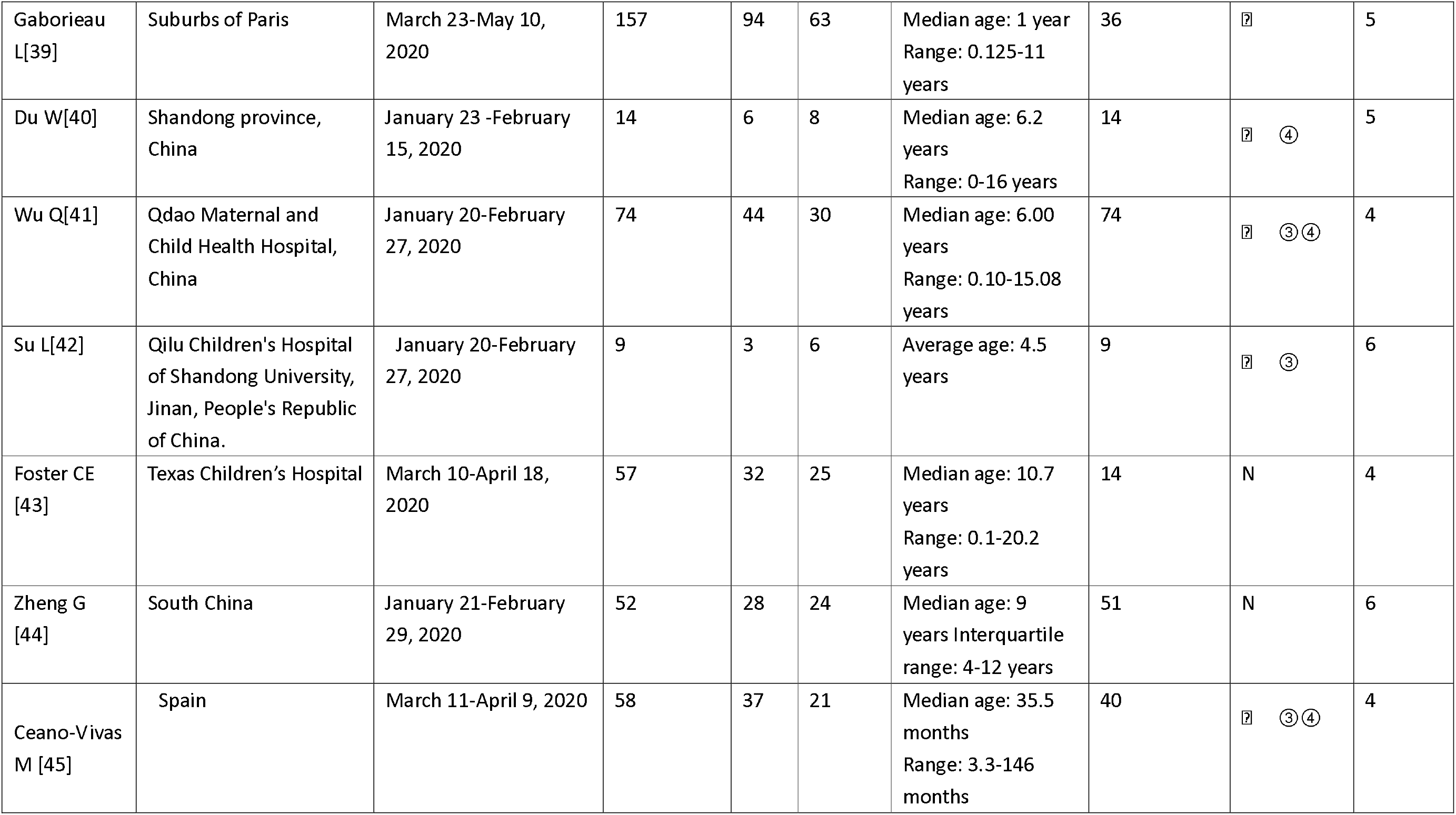
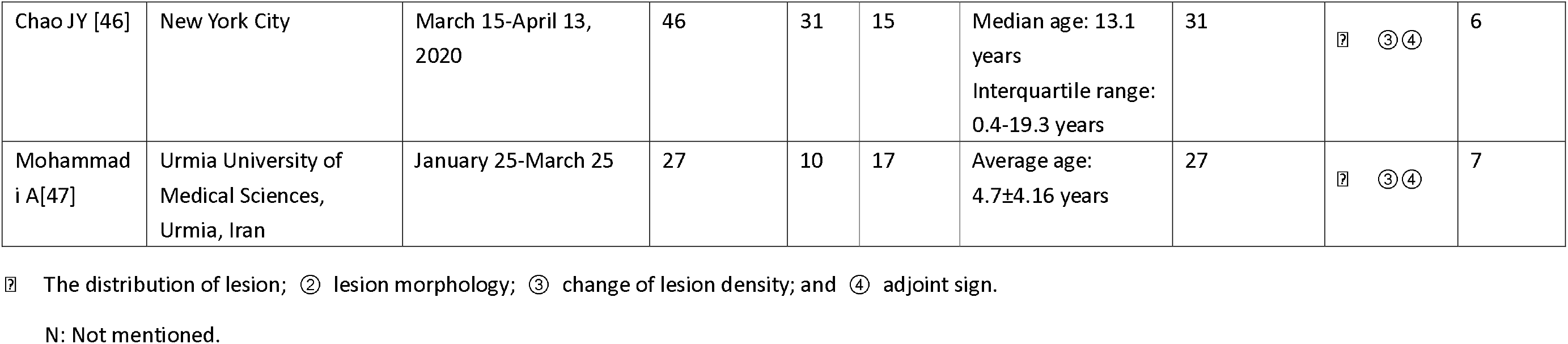
Basic fratures of the included studies

### 3.3 Meta-Analysis Results(Table 2)

**Table 2:** Summary of the meta-analysis results on CT image features CT: computed tomography.

| Outcome indicators | Number of included studies | Sample size | heterogeneity |  | Meta-analysis results |  |
| --- | --- | --- | --- | --- | --- | --- |
|  |  |  | *P-value* | *I*<sup>2</sup> | R% (95% confidence interval) | *P-value* |
| **Distribution of lesions** |  |  |  |  |  |  |
| Rate of abnormal CT findings | 37 | 1130 | 0 | 85.10% | 63.2 % (55.8-70.6) | 0 |
| Multiple lung lobes lesions | 12 | 184 | 0.768 | 0 | 65.1% (58.8-72.2) | 0 |
| Bilateral lung lesions | 25 | 300 | 0.169 | 22.80% | 61.5% (55.1-67.9) | 0 |
| **Lesion density** |  |  |  |  |  |  |
| Crazy-paving pattern | 10 | 183 | 0 | 0 | 6.15% (1.5-10.8) | 0.01 |
| Ground-glass opacities | 30 | 780 | 0 | 88.50% | 39.5% (30.7-48.3) | 0 |
| Air bronchogram sign | 7 | 121 | 0.01 | 0.00% | 11.2% (5.5-16.9) | 0 |
| Consolidation | 24 | 594 | 0 | 90.5 | 24.1% (15.9-32.4) | 0 |
| **Shape of lesions** |  |  |  |  |  |  |
| Nodules | 9 | 160 | 0.014 | 58.10% | 25.7% (15.6-35.9) | 0 |
| Patchy | 13 | 323 | 0 | 85.00% | 36.8% (23.5-50.1) | 0 |
| Cord-like | 6 | 129 | 0.173 | 37.20% | 9.7% (2.4-17.0) | 0.009 |
| Halo sign | 7 | 124 | 0.01 | 77.30% | 24.8% (8.5-41.2) | 0.003 |
| **Adjoint sign** |  |  |  |  |  |  |
| Pleural effusion | 5 | 239 | 0.05 | 70.20% | 9.1% (2.5-15.7) | 0.07 |

CT: computed tomography.

#### 3.3.1 Distribution of lesions: Meta-analysis of the random-effect model showed that the detection rate of abnormal lung CT findings was 63.2% (95% CI: 55.8–70.6%), bilateral lung lesions was 61.5% (95% CI: 58.8–72.2%), and multiple lung lobes lesions was 65.1% (95% CI: 55.1–67.9%)

#### 3.3.2 Lesion density

Meta-analysis of the random-effect model showed that the prevalence of nodules was 25.7% (95% Cl: 15.6–35.9%), patchy shadows was 36.8% (95% Cl: 23.5–50.1%), cord-like pattern was 9.7% (95% Cl: 2.4–17%), and halo signs was 24.8% (95% CI: 8.5–10.8%).

#### 3.3.3 Lesion shape

Meta-analysis of the random-effect model showed that the prevalence of a crazy-paving pattern was 6.1% (95% Cl: 1.5–10.8%), ground-glass opacities was 39.5% (95% CI: 30.7–48.3%), air bronchogram signs was 11.2% (95% CI: 5.5–16.98%), and consolidation was 24.1% (95% CI: 15.9–41.2%).

#### 3.3.4 Adjoint sign: Meta-analysis of the random-effect model showed that the prevalence of pleural effusion was 9.1% (95% CI: 2.5–15.7%). Two articles in this study reported cases of white lung [11, 17] and one article reported a case of pulmonary bullae [15],A study outside China reported 2 cases of pneumothorax[38]

#### 3.3.5 Subgroup analysis

The heterogeneity of this study was large. To explore the source of heterogeneity, the study was classified according to the region where the study took place (China and non-China) and grouped by the rate of abnormal pulmonary CT findings and ground-glass sample index. The results of each subgroup were consistent with the overall results, and there was no significant difference between the heterogeneity of each subgroup and the whole group, indicating that regional differences were not the main source of heterogeneity (Fig. 2 and 3).

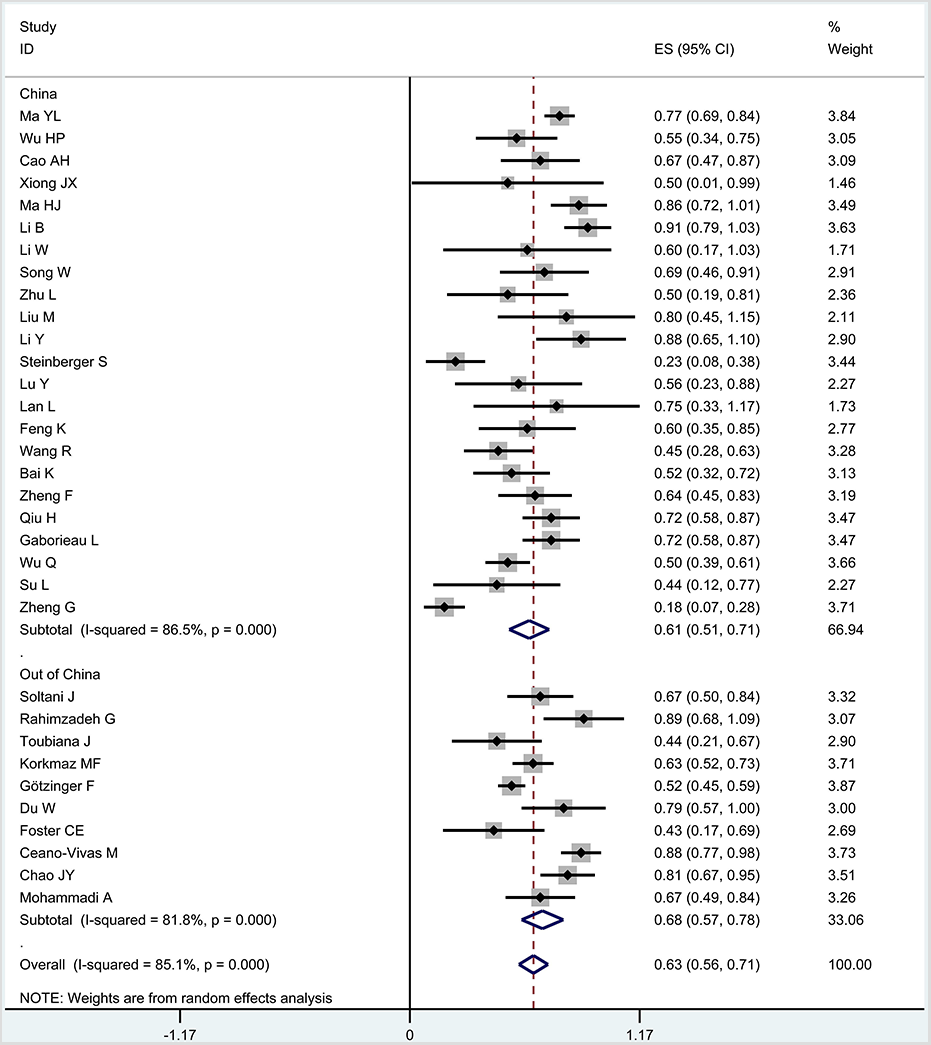

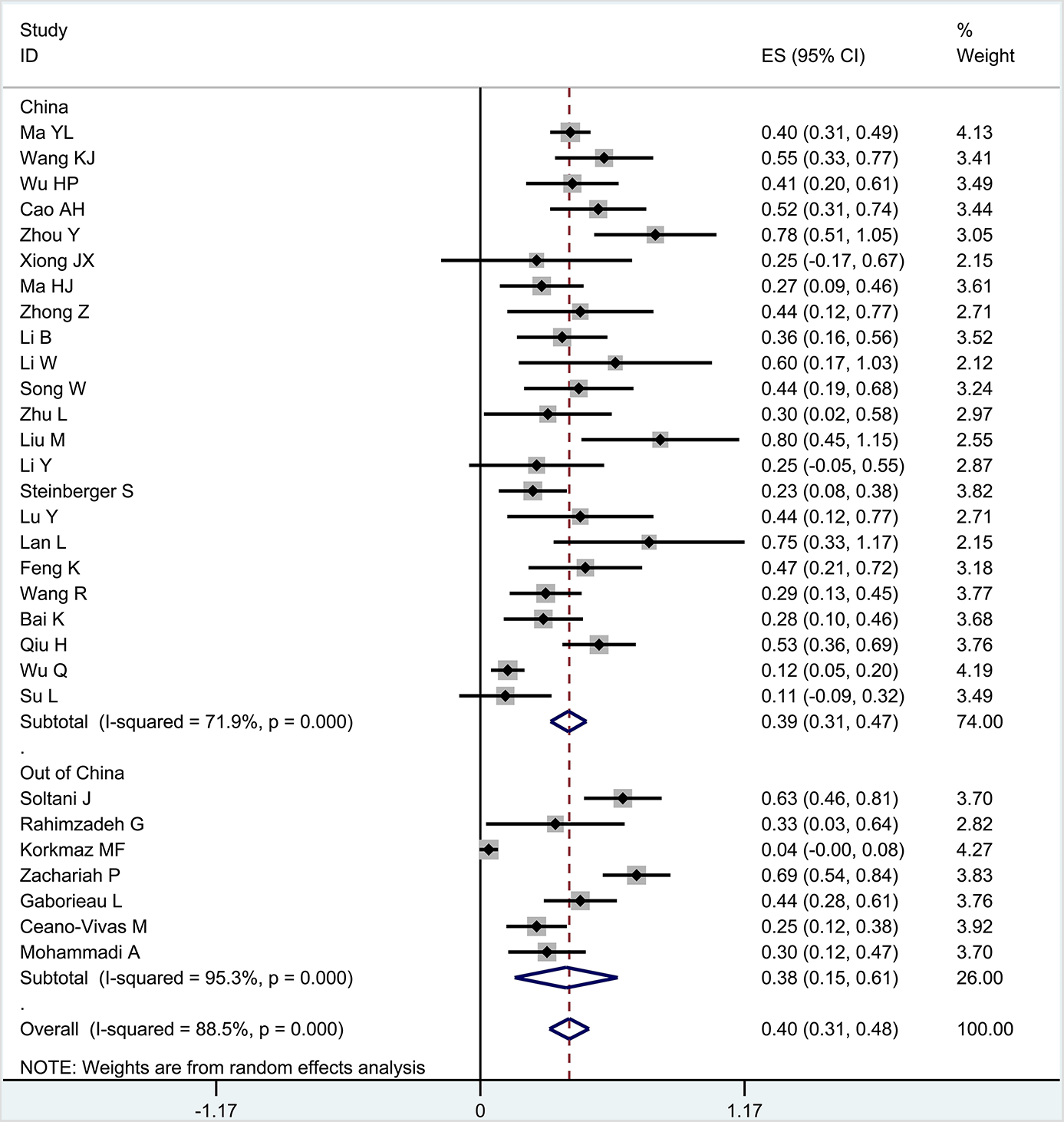

#### 3.4 Publication Bias

A funnel plot was drawn for the meta-analysis of abnormal lung CT indicators, and the results showed that the distribution at the left and right of each study point was symmetrical (Fig. 4), but the funnel plot of ground-glass indicators was asymmetric (Fig. 5), indicating that publication bias may exist in this study.

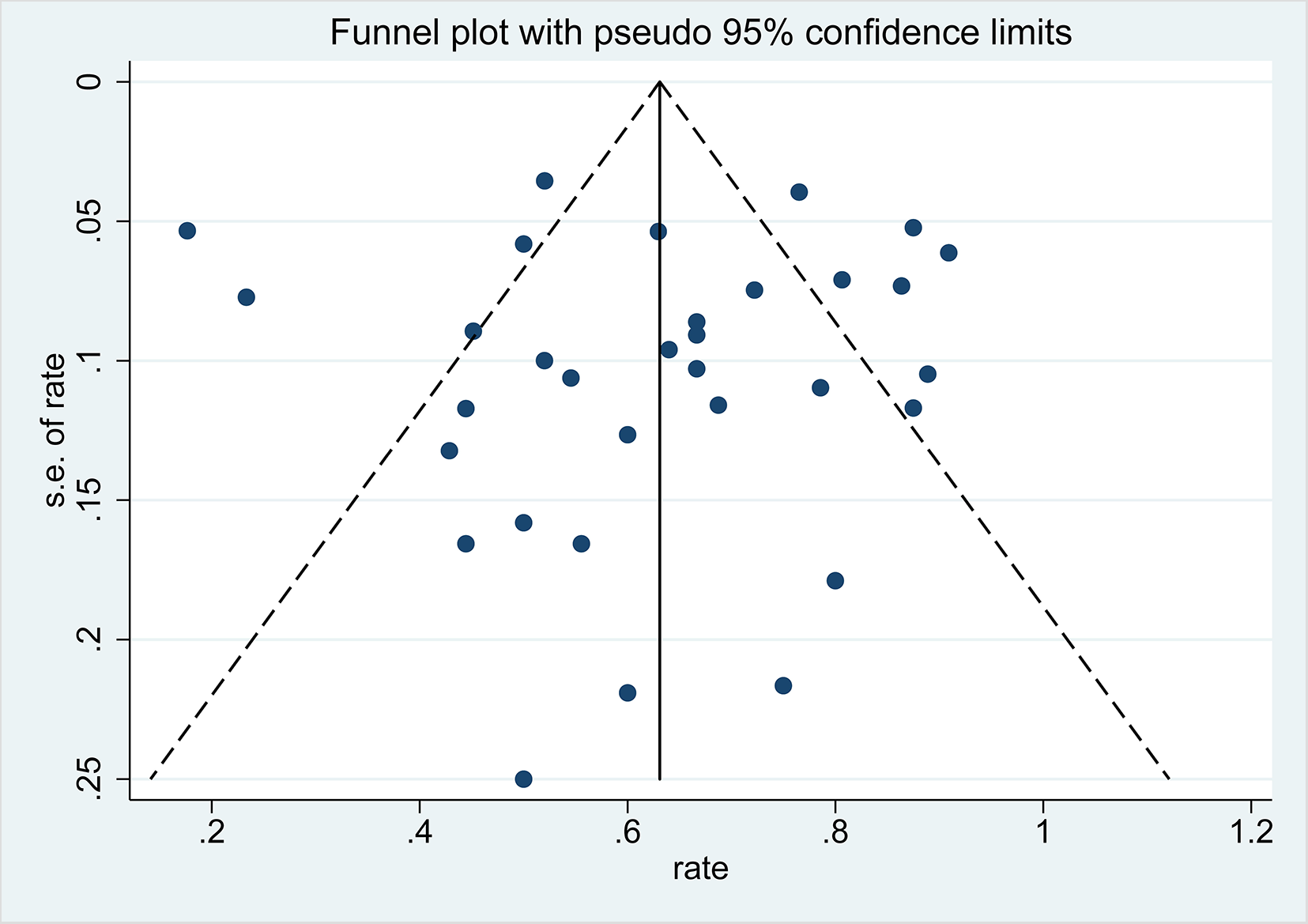

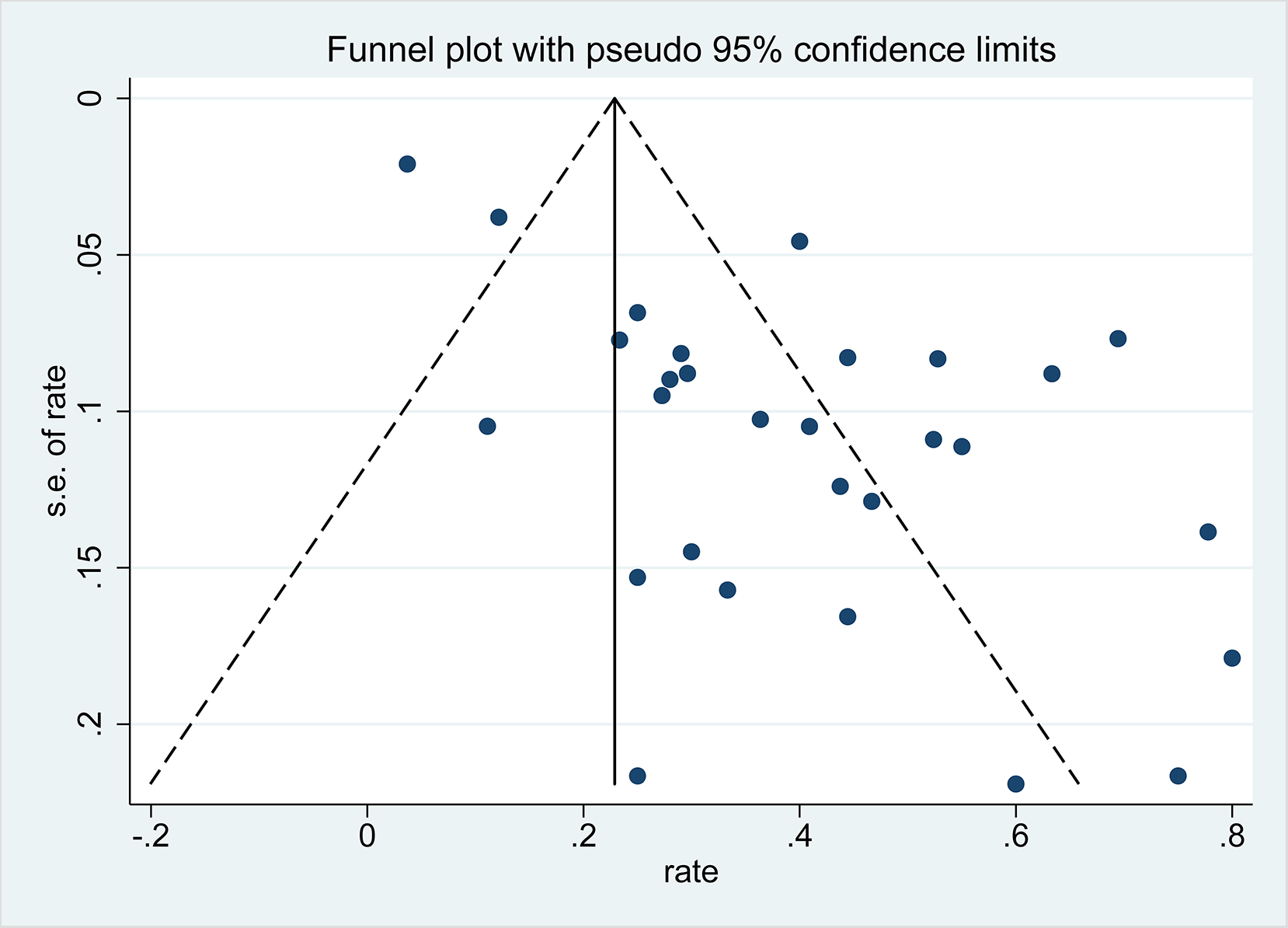

## 2. Discussion

Chest CT examination is a method used to diagnose COVID-19. CT manifestations of COVID-19 in adults mainly include patchy and segmental ground-glass density shadows in one or both lungs, or nodular shadows with surrounding ground-glass density shadows. It is mainly distributed around the vascular bundle, peripheral dorsal subpleural lung, and the lower lobes of both lungs, with the long axis parallel to the pleura. Air bronchogram and paving stone signs, among others, can also be observed [48]. In a systemic review, including 4121 cases of adult COVID-19 patients, it was found that the prevalence of typical ground-glass opacity was 68.1% (95% CI: 56.9–78.2%) and that most patients had bilateral lung involvement (73.8%; 95% CI: 65.9–81.1%) or lesions in multiple lung lobes (67.3%; 95% CI: 54.8–78.7%) [49]. Compared to adults, the current study found that the detection rate of abnormal CT findings in children was only 63.2% (95% CI: 55.8–70.6%), and the subgroup analysis showed that the basal rates of abnormal CT findings in and outside of China were 61% and 67.8%, respectively. While the incidence of typical ground-glass opacities was 39.5% (95% CI: 30.7–48.3%), the subgroup analysis revealed that the incidences in and outside of China were 39.2% and 38.1%, respectively.

Generally, the pulmonary CT scans of children do not show typical findings as that of adults. As for the CT examinations of asymptomatic children, Chinese scholar Lan [50] analyzed and found four cases with CT findings. Thin-section CT revealed abnormalities in three patients, and one patient did not present with any abnormal CT findings. Unilateral lung involvement was observed in two patients, and one patient showed bilateral lung involvement. In total, five small lesions were identified, including ground-glass opacity (n = 4) and consolidation (n = 1). All lesions had ill-defined margins with peripheral distribution and a predilection for the lower lobes. At present, there is no large-scale study on the differences of lung CT findings between asymptomatic and symptomatic children.

Generally, the symptoms of children are relatively mild. In the current study, two cases of white lung were reported from Wuhan Children’s Hospital;one article reported a case of pulmonary bullae [15],A study outside China reported 2 cases of pneumothorax[38]. The symptoms of children are milder than those of adults, which may be related to differences in the ACE2 receptor. ACE2 is an important binding receptor for SARS-CoV-2 [6,11]. The ACE2 receptor is still not quite mature in children and it also has reduced function than that in adults, making them less susceptible to SARS-CoV-2 binding. This also leads to reduced SARS-CoV-2 load. In addition, the immune system of children is still in the phase of development; therefore, the intensity of the immune response (cytokine storm) is not as strong as that in adults, which reduces the damage to the body [51].

Several limitations of our study need to be noted: ① Studies on children with COVID-19 are rare; there are 10 studies with less than 10 participants. Thus, the inspection efficiency may be insufficient. ② Most included studies were single-center studies; so, there may have been admission and selection biases. Most included studies were retrospective studies, which could not control for confounding factors. All these factors will affect the accuracy of the meta-analysis.

In conclusion, chest CT findings of children with COVID-19 are usually normal or slightly atypical; Thus, The CT findings show low sensitivity and specificity. Children diagnosed with COVID-19 are mainly diagnosed through reverse-transcription polymerase chain reaction. For children with a high suspicion of COVID-19, imaging examination shows no abnormalities and conclusions should be drawn cautiously.

## Data Availability

All data is viewed in the document

## Footnotes

## Contributors

JGW conceptualised and designed the study, contributed to the data selection, extraction and analysis, drafted the initial manuscript, reviewed and revised the manuscript and approved the final manuscript submitted. YFM managed the data, contributed to the data selection and extraction and did the meta-analyses, reviewed and revised drafts of the manuscript and approved the final manuscript submitted. YHS 、LCW、GBL、ML、QQQ contributed to the data selection and extraction, reviewed and revised drafts of the manuscript and approved the final manuscript submitted. All authors approved the final manuscript as submitted and agree to be accountable for all aspects of the work.

## Patient consent

Not required.

## Provenance and peer review

Not commissioned; externally peer reviewed.

## Data sharing statement

Extracted data are available upon request to the corresponding author.

